# Incidence and Risk Factors of Immediate Hypersensitivity Reactions and Immunisation Stress-related Responses with COVID-19 mRNA Vaccine

**DOI:** 10.1101/2022.01.12.22269134

**Authors:** Kazuo Imai, Fumika Tanaka, Shuichi Kawano, Kotoba Esaki, Junko Arakawa, Takashi Nishiyama, Soichiro Seno, Kosuke Hatanaka, Takao Sugiura, Yu Kodama, Seigo Yamada, Shinichiro Iwamoto, Shigeto Takesima, Nobujiro Abe, Chikako Kamae, Shigeaki Aono, Toshimitsu Ito, Tetsuo Yamamoto, Yasunori Mizuguchi

**Affiliations:** Self-Defense Forces Tokyo Large-scale Vaccination Centre, Tokyo, Japan; Self-Defense Forces Central Hospital, Department of Internal Medicine, Tokyo, Japan; Japan Ground Self-Defense Force, NBC Countermeasure Medical Unit, Tokyo, Japan; Self-Defense Forces Central Hospital, Department of Emergency Medicine, Tokyo, Japan; Japan Ground Self-Defense Force, Eastern Army Medical Unit, Tokyo, Japan; Self-Defense Forces Central Hospital, Department of Urology, Tokyo, Japan

**Author notes:** **Corresponding author** Kazuo Imai, M.D., Self-Defense Forces Central Hospital, Department of Internal Medicine, 1-2-24, Ikejiri, Setagaya-ku, Tokyo, Japan.

## Abstract

**Background:** With the implementation of mass vaccination campaigns against COVID-19, the safety of vaccine needs to be evaluated.

**Objective:** We aimed to assess the incidence and risk factors for immediate hypersensitivity reactions (IHSR) and immunisation stress-related responses (ISRR) with the Moderna COVID-19 vaccine.

**Methods:** This nested case-control study included recipients who received the Moderna vaccine at a mass vaccination centre, Japan. Recipients with IHSR and ISRR were designated as cases 1 and 2, respectively. Controls 1 and 2 were selected from recipients without IHSR or ISRR and matched (1:4) with cases 1 and cases 2, respectively. Conditional logistic regression analysis was used to identify risk factors associated with IHSR and ISRR.

**Results:** Of the 614,151 vaccine recipients who received 1,201,688 vaccine doses, 306 recipients (cases 1) and 2,478 recipients (cases 2) showed 318 events of IHSR and 2,558 events of ISRR, respectively. The incidence rates per million doses were estimated as – IHSR: 266 cases, ISRR: 2,129 cases, anaphylaxis: 2 cases, and vasovagal syncope: 72 cases. Risk factors associated with IHSR included female, asthma, atopic dermatitis, thyroid diseases, and history of allergy; for ISRR, they were younger age, female, asthma, thyroid diseases, mental disorders, and a history of allergy and vasovagal reflex.

**Conclusion:** In the mass vaccination settings, the Moderna vaccine can be used safely owing to the low incidence rates of IHSR and anaphylaxis. However, providers should beware of the occurrence of ISRR. Risk factor identification may contribute to the stratification of high-risk recipients for IHSR and ISRR.

## Introduction

Mass vaccination campaigns for COVID-19 are being implemented worldwide to overcome the ongoing global pandemic caused by SARS-CoV-2. A standardised two-dose regimen of the Pfizer-BioNTech^1^ and Moderna^2^ mRNA vaccines provided a high level of protection against COVID-19 and are widely used. To evaluate the safety of the mRNA vaccine, the acute and long-term adverse events following immunisation (AEFI) are being actively investigated by government agencies and the scientific community.

AEFI is grouped into five categories – vaccine product-related reaction containing immediate hypersensitivity reactions (IHSR), vaccine quality defect-related reaction, immunisation error-related reaction, immunisation stress-related responses (ISRR), and coincidental event.^3^ Investigations into AEFI which occur immediately after the injection, especially IHSR and ISRR, are particularly important to evaluate the safety of mass vaccine administration. Hitherto, the rate of anaphylaxis, which is an acute life-threatening and serious IHSR, to Pfizer-BioNTech and Moderna mRNA vaccines has been reported to be extremely low (2·5-23·9 cases per million doses);^4-6^ female gender and history of allergy were reported as prominent risk factors.^6,7^ In terms of ISRR, a high incidence rate of vasovagal syncope after receiving the COVID-19 vaccine (8·2 per 100,000 doses) has been reported, especially in females, adolescents, people with a mental disorder, or a history of vasovagal syncope.^8^

However, the majority of existing reports^4-8^ analysed a database of passive surveillance systems, such as the Vaccine Adverse Event Report System (VAERS)^9^ in the US. Because of well-documented limitations of passive surveillance systems, such as high inconsistencies in the report quality, and underreporting or biased reporting,^10^ previous studies mainly focused on the incidence rate and risk factors for anaphylaxis^4-7^ or vasovagal syncope^8^ as indicators of vaccine safety. However, there exists a knowledge gap regarding the incidence and risk factors for non-serious IHSR (e.g., skin, cardiac, gastrointestinal and respiratory symptoms except for anaphylaxis) and ISRR (e.g., symptoms and signs due to vasovagal reflex, panic attack and functional neurological disorders) that occur frequently and are crucial for clinicians who work in mass vaccination centres. Therefore, it is necessary to understand the detailed clinical characteristics of recipients with non-serious IHSR and ISRR to establish a safe mass vaccination system, and to identify the people who are at high risk for developing IHSR and ISRR.

At the Self-Defense Forces Tokyo Large-scale Vaccination Centre, Japan, a total of 1,201,688 Moderna COVID-19 vaccines have been administrated between May 24 and Sep 24, 2021, and active surveillance of AEFI was conducted. We also performed a detailed analysis of the clinical profile of all recipients both with and without IHSR and ISRR who received the Moderna COVID-19 vaccine to identify the incidence and risk factors of IHSR and ISRR.

## Materials and Methods

### Study design and participants

We conducted a nested case-control study at the Self-Defense Forces Tokyo Large-scale Vaccination Centre in Japan. An overview of the vaccination centre organisation and standard operating procedures of vaccine administration is shown in Supplementary methods 1. The adult vaccine recipients (≥18 years old) who received the Moderna COVID-19 vaccine between May 24 and Sep 24, 2021, were enrolled in this study. AEFI, which occurred during the stay of the recipient at the centre, was collected. Recipients who showed IHSR were designated as Case 1 and those who developed ISRR were designated as Case 2 (Figure 1). In case the recipients showed the same type of AEFI at both first and second doses, data for only the first dose were collected. If the recipient experienced two different types of AEFI at both first and second doses, each dose was selected. Vaccine recipients without AEFI were designated to controls. The period of vaccine administration and the number of vaccinations received were considered as potential confounders. Thus, controls 1 and controls 2 were matched (1:4) with cases 1 and 2 (Figure 1), respectively, based on the period of vaccine administration (May 24-June 23, June 24-July 23, July 24-Aug 23, and Aug 24-Sep 24) and the number of vaccinations received (first dose and second dose).

**Figure 1.**
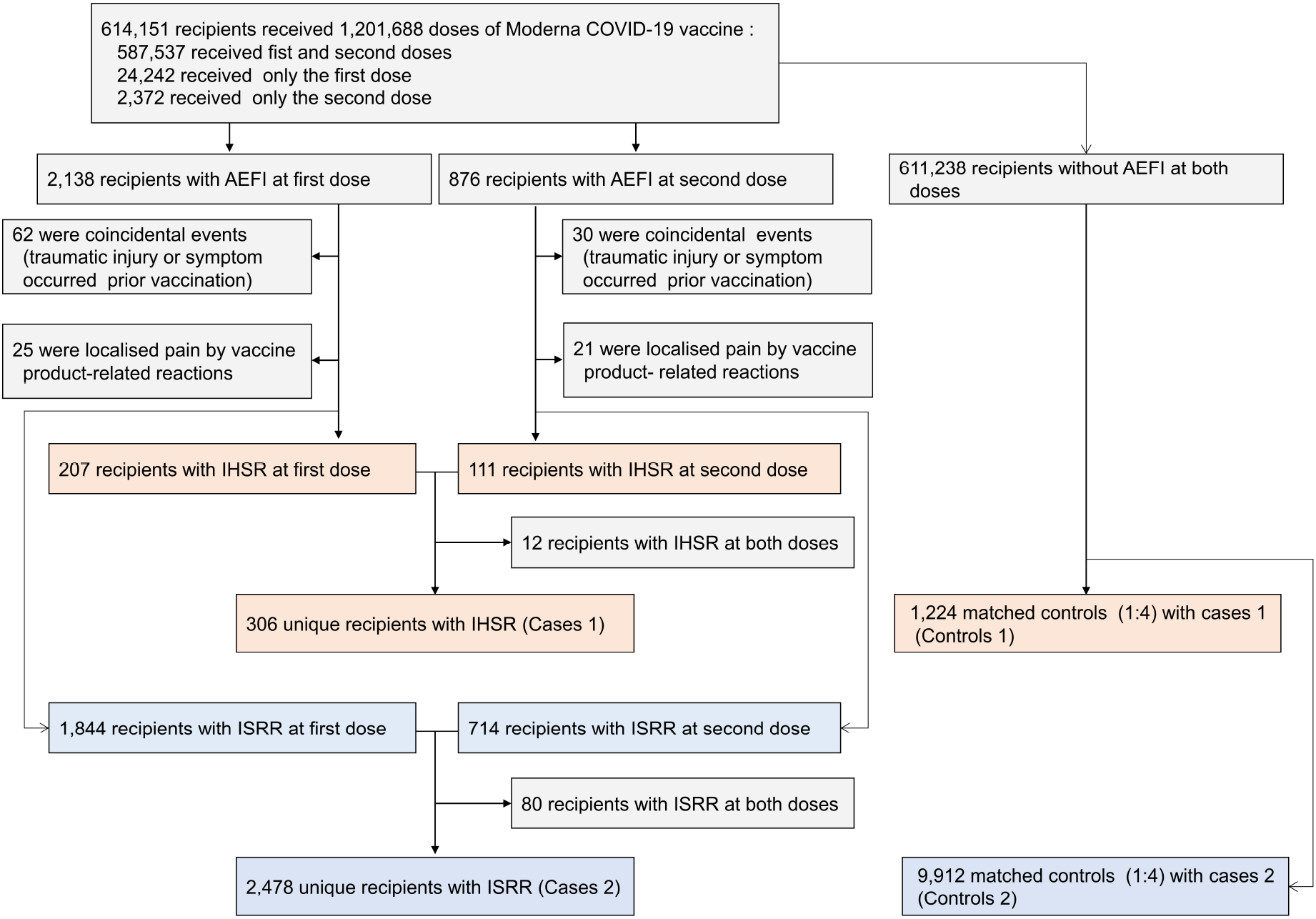
Flow diagram of vaccine recipients at the mass vaccination centre. AEFI; adverse events following immunisation, IHSR; immediate hypersensitivity reactions and ISRR; immunisation stress-related responses.

Baseline clinical characteristics (age, sex, comorbidities of hypertension, dyslipidaemia, diabetes, cardiovascular diseases, asthma, atopic dermatitis, thyroid diseases, malignancy and mental disorders, and history of allergic episodes for drugs and foods, and vasovagal episode) were collected using a pre-vaccination screening questionnaire for the COVID-19 vaccine distributed by The Ministry of Health, Labor and Welfare, Japan^11^ which was filled by the recipients before the injection and collected at each dose. Data for the date of vaccination and the number of doses received were collected using the in-house COVID-19 vaccine reservation and reception system (MRSO Inc, Tokyo, Japan). Next, all relevant clinical findings (symptoms, signs, the timing of onset of symptoms after the injection, medication received, if any, and the clinical outcome of requiring transportation to the hospitals and death) of the vaccine recipients with an AEFI were collected via medical records maintained in the first-aid rooms. During the study period, two trained physicians (KI and KE) and an emergency physician (FT) reviewed medical records to classify an AEFI daily.

This study was reviewed and approved by the Institutional Review Board of the Self-Defence Forces Central Hospital, Tokyo, Japan (Approval number: 03-006). Informed consent was obtained from all participants in the form of opt-out.

### Definitions

As defined by the World Health Organisation (WHO), ‘adverse event following immunisation (AEFI)’ was described as ‘‘any untoward medical occurrence which follows immunisation and which does not necessarily have a causal relationship with the use of the vaccine. The adverse event may be any unfavourable or unintended sign, an abnormal laboratory finding, a symptom, or a disease”.^3^ The standard criteria to classify AEFI are shown in Supplementary method 2.

Confirmatory anaphylaxis was diagnosed based on the Brighton Criteria.^12^ Grade 3 hypertension was defined as systolic blood pressure >180 mmHg and/or diastolic blood pressure >110 mmHg.^13^

### Statistical analysis

We calculated the incidence rates and 95% confidence interval (CI) for AEFI using the number of vaccine doses administrated at a centre as the denominator. Categorical variables are presented as frequency and percentage (%) and were compared using a chi-squared test or Fisher’s exact test, as appropriate.

As reported by the existing literature that age, sex, multiple comorbidities, and a history of allergy or vasovagal reflex after vaccination increased the risk of IHSR to other drugs^6,7,14-16^ and ISRR^8,17^. Therefore, we also selected comparable characteristics – age, sex, the presence of comorbidities (hypertension, diabetes, dyslipidaemia, cardiovascular diseases, asthma, atopic dermatitis, thyroid diseases, malignancy, and mental disorders), and a history of allergy to drugs and foods or a vasovagal reflex as variables of interest.

All variables that may be potentially associated with an increased risk of IHSR or ISRR as observed by univariate analysis (*p* < 0·10) were further processed through multivariable models. The final model was selected using backward stepwise conditional logistic regression to minimalise the Akaike information criterion (AIC). All models included age and sex and were adjusted by the period of the vaccine administration and the number of vaccination doses. A two-sided *p* value of < 0·05 was considered statistically significant. Variables for which < 25% of the data were missing, values were imputed with the use of multiple imputations by fully conditional specification using mice package.^18^

Sensitivity analyses were performed to test the robustness of the results by changing the definitions of IHSR and ISRR within the case. The case definition was gradually narrowed down to eliminate possible misclassifications between IHSR and ISRR (case definition of IHSR-2 and 3 or ISRR-2 and 3). Matched controls for each case of the IHSR and ISRR group were extracted from controls 1 or controls 2, respectively (Supplementary figures 1 and 2). In sensitivity analyses, we excluded recipients with missing data (complete case analysis). Case definitions and the number of participants in each sensitivity analysis are shown in Supplementary tables 1-4. Final models selected in the initial analysis were evaluated by new cases and their controls. Sample size consideration is shown in Supplementary method 3. All statistical analyses were performed using R software (v 4.0.2; R Foundation for Statistical Computing, Vienna, Austria; http://www.R-project.org/).

## Results

### Baseline characteristics of participants

Between May 24 and Sep 24, 2021, 614,151 people received the Moderna vaccine at the study centre (587,537 received both first and second doses, 24,242 received only a first dose and 2,372 received a second dose). A total of 1,201,688 vaccine doses (611,779 and 589,909 for a first dose and a second dose, respectively) were administrated at the study centre. During the study period, 3,014 instances of AEFI were observed in 2,913 recipients – a total of 101 recipients showed AEFI twice, both at first and second doses. Based on the clinical symptoms and signs, 318 events of IHSR were observed in 306 recipients (11%, cases 1) and 2,558 events of ISRR in 2,478 recipients (85%, cases 2) (Figure 1). Among the 611,238 recipients without AEFI, 1,208 recipients were selected as matched controls 1 for cases 1 and 9,940 recipients as control 2 for cases 2 (Figure 1). The baseline characteristics of all participants were shown in Table 1.

**Table 1.** Baseline characteristics.

|  | Immediate hypersensitivity reactions |  |  | Immunisation stress-related responses |  |  |
| --- | --- | --- | --- | --- | --- | --- |
|  | Cases 1<br>N=306 | Controls 1<br>N=1,224 | *p* value | Cases 2<br>N=2,478 | Controls 2<br>N=9,912 | *p* value |
| **Demographic characteristic** |  |  |  |  |  |  |
| **Age, years** |  |  |  |  |  |  |
| > 65 years | 108 (35) | 467 (38) | 0.817 | 430 (20) | 2344 (26) | <0.001 |
| 51-65 years | 69 (23) | 258 (21) |  | 361 (13) | 1913 (18) |  |
| 36-50 years | 68 (22) | 264 (22) |  | 620 (26) | 2815 (29) |  |
| ≤ 35 years | 61 (20) | 235 (19) |  | 1067 (41) | 2840 (27) |  |
| **Sex** |  |  |  |  |  |  |
| Male | 63 (21) | 692 (57) | <0.001 | 827 (33) | 5475 (55) | <0.001 |
| Female | 243 (79) | 532 (44) |  | 1651 (67) | 4437 (45) |  |
| **Comorbidities** |  |  |  |  |  |  |
| Hypertension | 40 (13) | 156 (13) | 0.848 | 163 (7) | 899 (9) | <0.001 |
| Dyslipidaemia | 17 (6) | 58 (5) | 0.552 | 89 (4) | 406 (4) | 0.303 |
| Diabetes | 12 (4) | 59 (5) | 0.647 | 55 (2) | 351 (4) | 0.002 |
| Cardiovascular diseases | 5 (2) | 40 (3) | 0.182 | 57 (2) | 197 (2) | 0.284 |
| Asthma | 24 (8) | 17 (1) | <0.001 | 96 (4) | 123 (1) | <0.001 |
| Atopic dermatitis | 5 (2) | 3 (0) | 0.010 | 21 (1) | 36 (0) | 0.002 |
| Thyroid diseases | 15 (5) | 13 (1) | <0.001 | 50 (2) | 89 (1) | <0.001 |
| Malignancy | 6 (2) | 21 (2) | 0.807 | 26 (1) | 90 (1) | 0.476 |
| Mental disorders | 7 (2) | 20 (2) | 0.463 | 113 (5) | 127 (1) | <0.001 |
| Missing data of comorbidities | 4 (1) | 0 | .. | 33 (1) | 0 | .. |
| **History** |  |  |  |  |  |  |
| Allergic episodes for drugs | 86 (28) | 41 (3) | <0.001 | 320 (13) | 303 (3) | <0.001 |
| Allergic episodes for foods | 80 (26) | 33 (3) | <0.001 | 308 (12) | 327 (3) | <0.001 |
| Vasovagal episode | 9 (3) | 10 (1) | 0.006 | 246 (10) | 136 (1) | <0.001 |
| Missing data of histories | 6 (2) | 0 | .. | 45 (2) | 10 (0) | .. |
| **Number of vaccine received** |  |  |  |  |  |  |
| First | 207 (68) | 828 (68) | 1.000 | 1844 (74) | 7376 (74) | 1.000 |
| Second | 99 (32) | 396 (32) |  | 634 (26) | 2536 (26) |  |
| **Period** |  |  |  |  |  |  |
| May 24-June 23 | 98 (32) | 392 (32) | 1.000 | 486 (20) | 1944 (20) | 1.000 |
| June 24-July 23 | 58 (19) | 232 (19) |  | 323 (13) | 1292 (13) |  |
| July 24-Aug 23 | 89 (29) | 356 (29) |  | 1071 (43) | 4284 (43) |  |
| Aug 24-Sep 24 | 61 (19) | 244 (20) |  | 598 (24) | 2392 (24) |  |

447 Data are n (%).

### Risk factor analysis

In the univariable analysis, we observed that recipients with ISRR were significantly younger (*p* < 0·001) than their controls (cases 2 vs controls 2), but no significant difference was found in the IHSR group (cases 1 vs controls 1). The proportion of females were significantly higher (*p* < 0·001) in recipients with both IHSR and ISRR than in their respective controls. The variables potentially associated with the IHSR group included asthma, atopic dermatitis, thyroid diseases, history of allergy to drugs and foods, and history of vasovagal reflex; those potentially associated with ISRR group included hypertension, diabetes, asthma, atopic dermatitis, thyroid diseases, mental disorders, history of allergy to drugs and foods, and history of vasovagal reflex (*p* < 0·100) (Table 1).

In the multivariable conditional logistic regression analysis, the variables significantly associated with an increased risk of IHSR included female gender, asthma, atopic dermatitis, thyroid diseases, and history of allergy to drugs and foods (Figure 2A), of which, history of allergy to drugs (odds ratio, OR: 13·32 [95% CI: 7·57–23·44]) and foods (OR: 11·80 [95% CI: 7·04–19·80]) had the strongest association for an increased risk of IHSR (Figure 2A). Similarly, in recipients who developed ISRR, younger age (≤ 65 years), female gender, asthma, mental disorders, history of allergy to drugs and foods, and history of vasovagal reflex were significantly associated with an increased risk (Figure 2B). Younger the recipient’s age, the greater the risk of developing ISRR.

**Figure 2.**
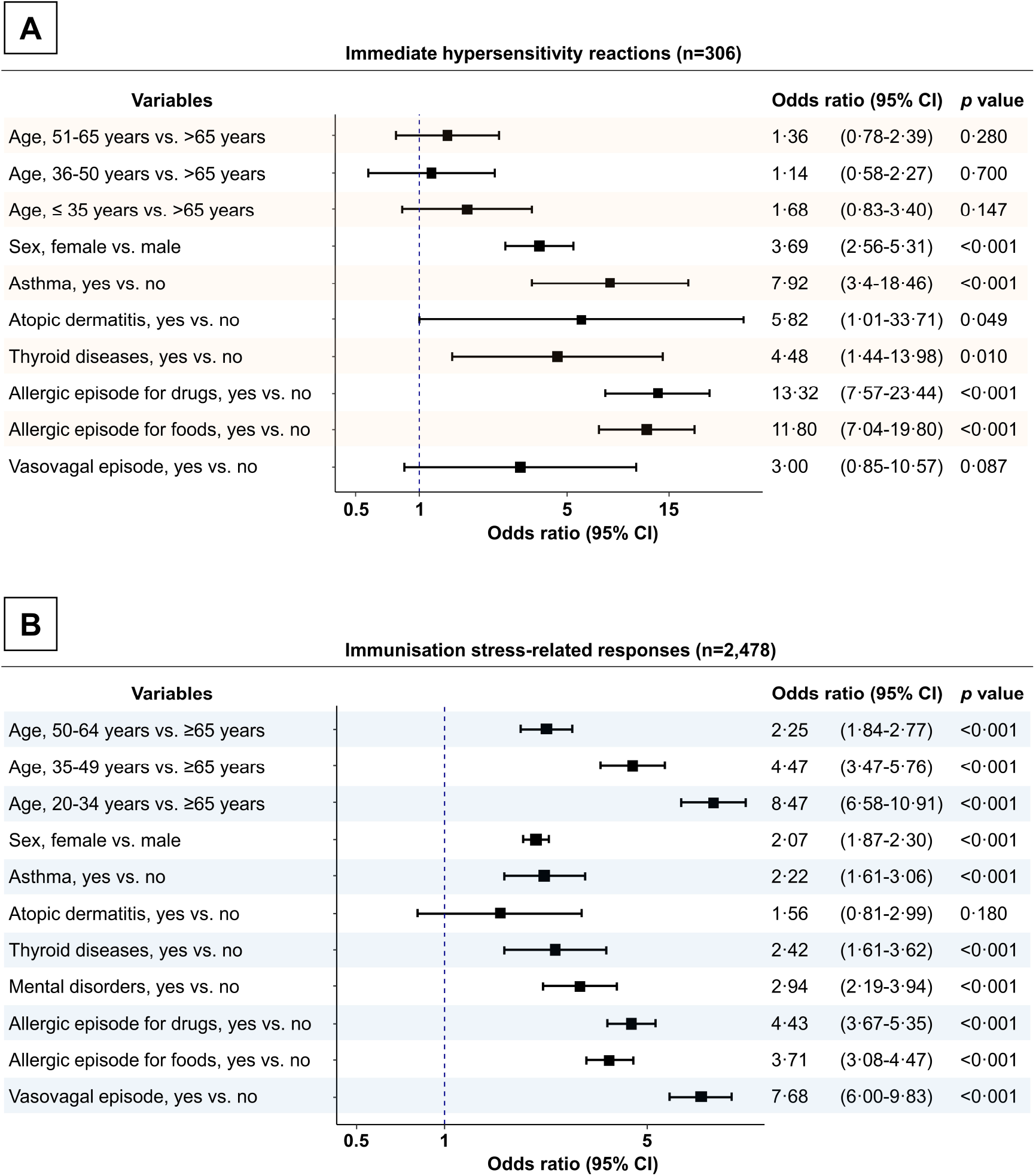
Risk factors associated with immediate hypersensitivity reactions and immunisation stress-related responses to the Moderna COVID-19 vaccine. (A) Forest plot showing the odds ratio for an increased risk of immediate hypersensitivity reactions and (B) immunisation stress-related responses using multivariable analysis of conditional logistic regression analysis. Plots and horizontal lines indicate estimated odds ratio and 95% confidence intervals, respectively.

### Sensitivity analysis

According to the new case definition, 284 cases (IHSR-2) and 188 cases (IHSR-3) were selected from the case 1 group and were compared to 1,136 and 752 matched-controls, respectively. Likewise, 2,304 cases (ISRR-2) and 2,129 cases (ISRR-3) were selected from cases 2 to compare with 9,208 and 8,516 matched controls selected from controls 2, respectively (Supplementary figures 3 and 4). We observed that changing the case definitions of IHSR and ISRR did not change the study findings, although the association of atopic dermatitis for an increased risk of IHSR did not reach statistical significance as per the new case definition.

### Incidence rates of IHSR and ISRR

Of the 318 IHSR events, two events were classified as anaphylaxis according to the Brighton Criteria (one event at the first dose – level 2-2, and another at the second dose – level 3). Of the 2,558 IHSR events, 86 events of vasovagal syncope were observed. Overall, the incidence rate per million doses of AEFI in the 1,201,688 vaccine doses administered was estimated as follows– IHSR: 266 cases (95% CI; 236-295 cases), ISRR: 2,129 cases (95% CI; 2047-2212 cases), anaphylaxis: 2 cases (95% CI; 0·2-6 cases), and vasovagal syncope: 72 cases (95% CI; 57-88 cases) (Figure 3A and Supplementary table 5). The incidence rate of AEFI at the first dose, except for anaphylaxis, was significantly higher than at the second dose (*p* < 0·001) (Supplementary table 5).

### Symptoms and signs of vaccine recipients with IHSR and ISRR

Vertigo, malaise, and numbness or loss of sensation in part of the body were the most common clinical symptoms (Figure 3B and Supplementary tables 6 and 7). In the IHSR events, any types of rash and pruritus were the most common symptoms. Syncope was observed in the ISRR events only, and all of these cases were diagnosed with vasovagal syncope. Hypotension and bradycardia due to vasovagal reflex were the most common vital sign abnormalities in the ISRR events, whereas grade 3 hypertension was the most common vital sign abnormality in the IHSR events.

**Figure 3.**
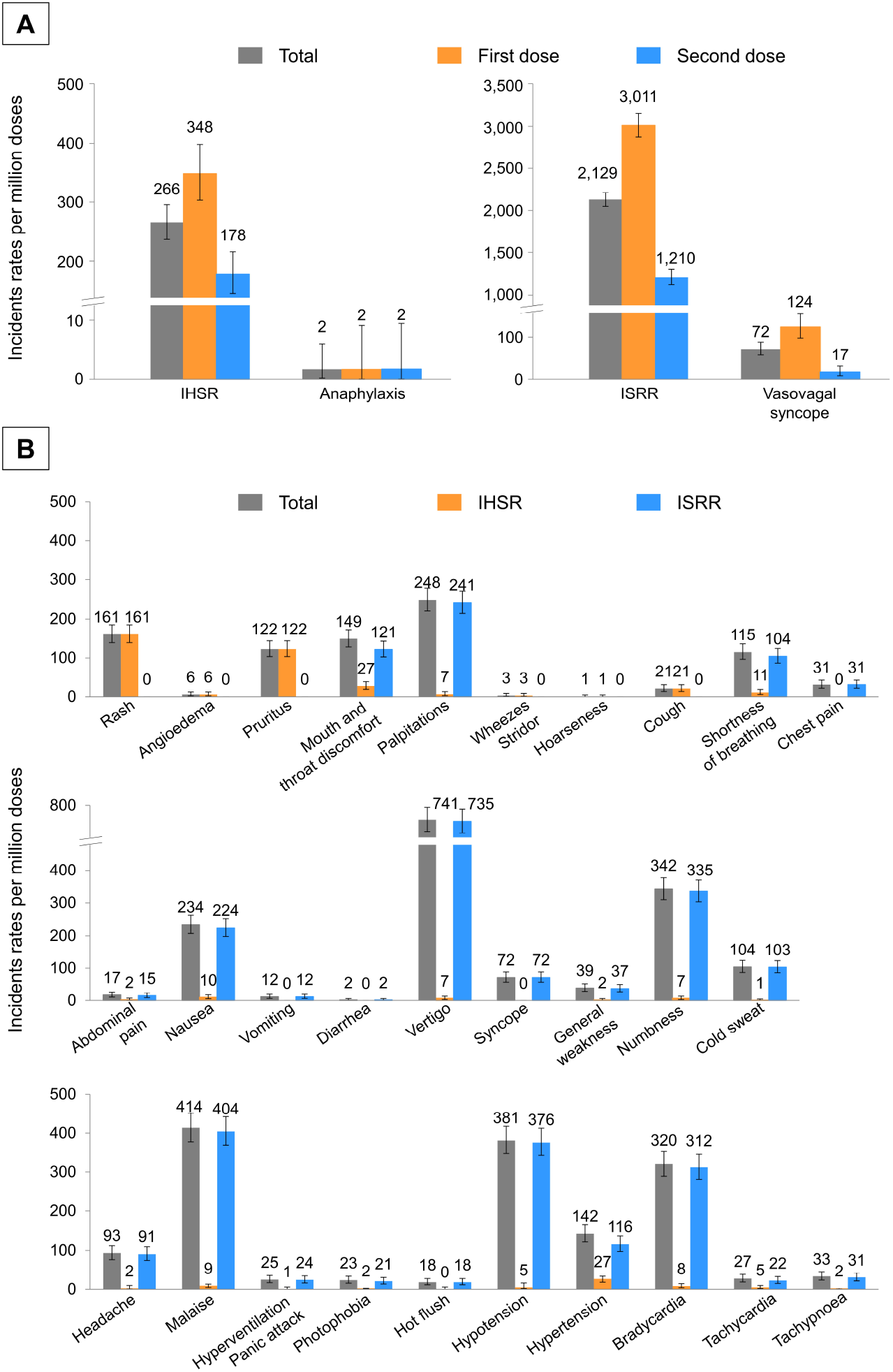
Incidence rates of adverse events following immunisation to Moderna COVID-19 vaccine. (A) Bar plot showing the incidence rates of immediate hypersensitivity reactions (IHSR) and immunisation stress-related responses (ISRR) and of (B) clinical symptoms and signs. The incidence rates were estimated using vaccine doses administrated as the denominator. Error bars indicate 95% confidence intervals.

Symptom onset within 15 minutes of the vaccination was recorded in 179 (56%) of 318 events of IHSR and 1,928 (75%) of 2,558 events of ISRR, whereas, in 294 (93%) of the 318 events of IHSR and 2,450 (96%) of the 2,558 events of ISRR, the symptoms appeared within 30 minutes of vaccination (Table 2). Epinephrine was used for seven events with a clinical diagnosis of severe IHSR at the first-aid rooms. Erroneous administration of epinephrine for hypotension due to vasovagal reflex was reported for two events in the ISRR events. A total of 75 events (3%) required additional treatment in neighbouring hospitals. Twenty-seven events (9%) required additional treatment at neighbouring hospitals in the IHSR event, whereas 48 events (2%) required additional examination at neighbouring hospitals in the ISRR events including 14 events of persistent grade 3 hypertension and 17 events of neurological disorders (one or more symptoms of general weakness, numbness, loss of sensation, and movement disorders). In the IHSR events, 287 events (90%) were self-limiting requiring no medication or treatment.

**Table 2.** Descriptive characteristics of vaccine recipients with acute adverse events following immunisation.

|  | No. of total events<br>N=2,876 | No. of events of<br>immediate<br>hypersensitivity<br>reactions<br>N=318 | No. of events of<br>immunisation<br>stress-related<br>responses<br>N=2,558 |
| --- | --- | --- | --- |
| **Onset of initial symptoms/signs** |  |  |  |
| ≤15 min | 2107 (73) | 179 (56) | 1928 (75) |
| ≤30 min | 2744 (95) | 294 (93) | 2450 (96) |
| >30 min | 71 (3) | 17 (5) | 54 (2) |
| Missing | 61 | 7 | 54 |
| **Medications at a centre** |  |  |  |
| Epinephrine | 9 (0) | 7 (2) | 2 (0) |
| Antihistamine | 3 (0) | 3 (1) | 0 |
| Corticosteroid | 1 (0) | 1 (0) | 0 |
| Both of two | 1 (0) | 1 (0) | 0 |
| **Outcome** |  |  |  |
| Transported to the hospitals | 75 (3) | 27 (9) | 48 (2) |
| Recovered at a centre | 2801 (97) | 291 (92) | 2510 (98) |
| Recovered with medication | 6 (0) | 4 (1) | 2 (0) |
| Recovered without medication | 2795 (97) | 287 (90) | 2508 (98) |
| Death | 0 | 0 | 0 |

Data are n (%)

## Discussion

This single-centre nested case-control study provides an outline of the incidence rates and risk factors for developing IHSR and ISRR among recipients of the Moderna COVID-19 vaccine in Japan. We conducted active surveillance at our centre to document the clinical findings of all recipients who developed an AEFI regardless of the severity. Notably, more than 80% of all instances AEFI were classified as ISRR, instead of IHSR, and the incidence rates of both IHSR and ISRR were significantly higher after the first dose compared to the second one, however, the overall incidence of AEFI was very low at both first and second doses. By comparing clinical characteristics between recipients with and without AEFI, we identified several risk factors associated with the development of IHSR and ISRR.

We observed that the incidence rate of IHSR was very low, approximately 266 cases per million doses (0·03%) of all recipients, at both first and second doses, which is significantly different from the estimated incidence reported previously. Blumenthal et al. conducted a questionnaire-based study and described that the incidence rate of IHSRs within three days after the injection was 2·1% out of 64,900 health care employees who received their first dose of Pfizer-BioNTech and Moderna COVID-19 mRNA vaccines.^19^ Contrarily, Myles and colleagues conducted real-time surveillance by an allergist at a mass vaccination centre and reported an incidence rate of 0·12% of IHSR among the 14,655 vaccine recipients.^20^ Our results are in line with those reported by Myles et al,^20^. Likewise, the anaphylaxis rate was also extremely low in our study (2 cases per million doses), which is consistent with the existing data reports for the US and UK.^4-6^

The mechanisms of IHSR following the Moderna vaccine administration are not completely clarified; however, PEG-2000 is the identified candidate allergen.^21^ Currently, only the female gender and history of allergy are known factors associated with an increased risk of anaphylaxis to mRNA vaccines. Shimabukuro et al. reported that anaphylaxis was more frequently observed in females than males.^6^ Similarly, Desai et al. documented that people with a history of allergy and anaphylaxis had a 2-7 times higher incidence of anaphylaxis following vaccination compared to people without any history of allergy.^7^ Our findings were comparable to these studies regarding the risk factors for IHSR. Additionally, we identified comorbidities (asthma, atopic dermatitis, and thyroid diseases) that were associated with a greater risk of IHSR. Asthma and atopic dermatitis are established risk factors for IHSR to several drugs.^16^ Thus, it seems that populations with atopic dermatitis and asthma are predisposed to develop IHSR to drugs, including mRNA vaccines, but not the response may not be specific to mRNA vaccine components, such as PEG-2000. Thyroid diseases was also identified as a risk factor for IHSR to contrast media,^15^ but not to other common drugs. Probably, people with thyroid diseases have a higher PEG-sensitisation rate than the general population.

It is known that ISRR is caused by anxiety and fear about injection, needles, vaccine components, adverse events, or pre-existing conditions.^17^ The incidence rate of ISRR in our study was low but not enough to be ignored (2,129 cases per million doses: 0·21%). A notable finding was the significantly high incidence rate of vasovagal syncope (72 cases per million doses), although people who had a history of vasovagal reflex were screened and were administrated the vaccine in a lying position. Hause et al. also reported a high incidence of vasovagal syncope following the J&J/Janssen COVID-19 vaccine, a viral vector vaccine, estimated at 8·2 per 100,000 doses in mass vaccination centres in the US.^8^ Strikingly, the incidence rate of vasovagal syncope after COVID-19 vaccines was significantly higher than the influenza vaccine (0·05 per 100,000 doses),^8^ and similar to that of the quadrivalent human papillomavirus vaccine (7·8 cases per 100,000 doses).^22^ In addition, we found that several clinical symptoms and signs resulted from ISRR, especially, a type of neurological symptoms, known as the functional neurological disorders (FND),^23^ that were difficult to assess as caused by psychological or organic factors in the setting of mass vaccination. Indeed, 2% of recipients with ISRR were transported to hospitals, and one-third of these patients showed neurological symptoms most likely due to FNDs, although more careful evaluations are needed for the diagnosis. Further studies with active surveillance are needed for a better understanding of the incidence of FNDs and to take appropriate mitigating measures at mass vaccination centres.

For effective implementation of precautionary measures, it is important to first identify individuals with a high risk of ISRR.^17^ In general, adolescence, female gender, mental disorders, and history of vasovagal reflex were considered as the risk factors for ISRR.^8,17^ Our findings reaffirm that these risk factors increase the chances of developing an ISRR after mRNA vaccines. Additionally, specific comorbidities (asthma and thyroid diseases) and a history of allergy were identified as additional risk factors of ISRR. There are several possibilities regarding these associations. The presence of comorbidities and a history of allergy may provoke strong anxiety and fear about the allergic adverse events and the effects of vaccination on comorbidities, especially during the first dose. The present scenario that this mRNA vaccine is a novel type of vaccine for infectious diseases may increase the associated fear and anxiety. Also, asthma^24^ and thyroid diseases^25^ increase the risk of mental disorders by 1·5 times and 2·3-3·5 times, respectively, and undiagnosed or underreported mental disorders may increase the risk of ISRR. Therefore, at mass vaccination centres, providers should be aware that a greater proportion of recipients are predisposed to ISRR after vaccination compared to the known high-risk cases.

Several symptoms, especially respiratory and gastrointestinal, are overlapped between IHSR and ISRR. To check for potential misclassification between IHSR and ISRR, we performed sensitivity analyses by narrowing the case definitions to reduce the effect of misclassification. The risk factors for IHSR and ISRR were consistent in our sensitivity analysis even while using narrowed case definitions, except for atopic dermatitis for IHSR with low prevalence, suggesting there is little effect of misclassification.

There are several limitations to this study. First, a selection bias may limit the generalisability of our findings. This study was conducted at a single centre and a single country. ISRR can be affected by environmental factors of the vaccination centre and can occur in clusters or group settings.^26^ Therefore, the incidence rate of ISRR cannot be specified in a mass vaccination centre. Second, the possibility of underestimation of the incidence rate of IHSR and ISRR cannot be denied. The observation period after the administration of the vaccine was limited (mean observation time was 20 minutes: Supplementary method 1) due to the study design. Third, the sample size of the case with IHSR was smaller than planned due to the low incidence rate of IHSR in our study (Supplementary method 2). Therefore, our sample size of IHSR may not have statistical power to detect the factors that slightly increased the risk of IHSR with a low prevalence rate, such as atopic dermatitis, in the sensitivity analysis. We suggest that multicentre and multinational studies may complement the limitations of our study.

## Conclusion

The incidence rate of IHSR and anaphylaxis in our single-centre study was very low, suggesting that the Moderna COVID-19 vaccine can be used safely for mass vaccinations. However, healthcare providers need to take appropriate measures to prevent and respond adequately to the development of ISRR. The identified risk factors in the present study will be useful for the stratification of the high-risk recipients regarding IHSR and ISRR and will contribute to understanding the mechanisms of IHSR to mRNA vaccines.

## Declarations

### Ethics approval and consent to participate

This study was reviewed and approved by the Self-Defence Forces Central Hospital (Approval number: 03-006). Informed consent was obtained in the form of opt-out.

### Consent for publication

Not applicable.

### Availability of data and materials

The datasets analysed during the current study are available from the corresponding author on reasonable request.

## Acknowledgments

We thank all participants who worked at the Self-Defence Forces Tokyo Large-scale Vaccination Centre, Japan for supporting the investigation and Mr. Shingo Tamaki at the Nagasaki University School of Tropical Medicine and Global Health for their helpful advice with the statistical analysis.

## Authors’ contributions

KI, study conceptualisation; FT, KE, TN, SS, KH, TS, YK, SY, SI and ST, investigation; KI, FT, and KE, data curation; KI and KE, performing formal analysis; KI, SK, JA, NA, CK, and TY, manuscript drafting; TN, SS, KH, TS, YK, SY, SI, ST, and SA, manuscript revision; TI and YM, study supervision. All authors have read and approved the final manuscript.

