## Supplementary materials for "Incidence and Risk Factors of Immediate Hypersensitivity Reactions and Immunisation Stress-related Responses with COVID-19 mRNA Vaccine"

### Contents


| **Methods** | | **Page** |
|---|---|---|
| **Supplementary method 1** | The overview of The Self-Defense Forces Tokyo Large-scale Vaccination Centre in Japan | 2,3 |
| **Supplementary method 2** | The standard criteria to classify acute adverse events following immunisation | 4 |
| **Supplementary method 3** | Sample size calculation | 5 |
| **Tables** | | |
| **Supplementary table 1** | Case definition of immediate hypersensitivity reaction in sensitivity analyses | 6 |
| **Supplementary table 2** | Case definition of immunisation stress-related response in sensitivity analyses | 7 |
| **Supplementary table 3** | Participants of immediate hypersensitivity reactions groups in sensitivity analysis | 8 |
| **Supplementary table 4** | Participants of immunisation stress-related response groups in sensitivity analysis | 9 |
| **Supplementary table 5** | Estimated incidence rates of adverse events following immunisation | 10 |
| **Supplementary table 6** | Number of events of clinical symptoms and signs | 11,12 |
| **Supplementary table 7** | Incidence rates of events of clinical symptoms and signs | 13,14 |
| **Figures** | | |
| **Supplementary figure 1** | Flow diagram of immediate hypersensitivity reactions groups in sensitivity analysis | 15 |
| **Supplementary figure 2** | Flow diagram of immunisation stress-related response groups in sensitivity analysis | 15 |
| **Supplementary figure 3** | Multivariable conditional logistic regression for immediate hypersensitivity reactions group in sensitivity analysis | 16 |
| **Supplementary figure 4** | Multivariable conditional logistic regression for immunisation stress-related response group in sensitivity analysis | 17 |

### Supplementary method 1

### The overview of The Self-Defense Forces Tokyo Large-scale Vaccination Centre in Japan

**Location**

A center located in Otemachi, Chiyoda-ku, Tokyo, Japan.

**Floor layout**

1 F: Entrance and a first aid-room

2F: Vaccination floor and a first aid-room.

4F: Vaccination floor and a first aid-room.

7F: Vaccination floor and a first aid-room.

10F: Vaccination floor and a first aid-room.

Each first-aid room located close to each observation room, except for 1F.

**Dosing and Schedule**

Moderna COVID-19 vaccine was used in the centre. Vaccine was stored at -20°C before its use and then thawed to the required temperature. Within 6 hours after thawing, vaccine was administered intramuscularly (0·5. mL each). Standard vaccination interval between a first and a second dose was 28 days, and permissible interval was 21-42 days.

**Target population**

The centre initially targeted people aged over 65 years who lived in Kanto area, Japan, from May 24 to Jun 16, 2021, and then the target was expanded to people aged over 18 years who lived in Japan from Jun 17 to Sep 24, 2021.

**Exclusion criteria for vaccine administration**

According to the national guideline, exclusion criteria for vaccine administration in the centre was defined as the following; people who have a fever (body temperature > 37·5°C), acute serious illness that is treatable and anaphylaxis episode for polyethylene glycol (PEG).

**The number of administration doses**

The target number of administration doses in the centre was 10,000 doses per day. During May 24 to Sep 24, a total of 1,201,688 vaccine doses (611,779 and 589,909 for a first dose and a second dose, respectively) were administrated in the centre. The average of vaccination doses per day was 9,770 doses.

**Administration procedure**

1. All recipients need to fill all items in pre-vaccination screening questionnaire for COVID-19 vaccine distributed by The Ministry of Health, Labour and Welfare, before the injection of each first and second dose.

2. A nurse checked all items in pre-vaccination screening questionnaire. A nurse asks recipients detailed questions about their comorbidities, history of allergy, and history of vasovagal reflex after the injection or collecting blood. If a misstatement or omission was found, the recipient is requested to revise and fill all items.
3. A medical doctor conducted medical inquiries for recipients and checked all items in pre-vaccination screening questionnaire. The medical doctor determined the observation period after the injection based on the history of allergy. Observation period was 30 minutes for recipient who has a history of anaphylaxis for any causes and 15 minutes for other all recipients.
4. The trained nurse injected the vaccine intramuscularly for the recipients. Generally, a recipient was injected sitting on a chair. If a recipient had a history of vasovagal reflex, then the recipient was injected lying on the bed.
5. Centre staff collected pre-vaccination screening questionnaire. Recipients who received a first dose of vaccination got a reservation for a second dose.
6. Recipient was observed for his or her condition in the observation room according to their observation period. The average time of staying time from the injection to leaving the centre was 20 min. If the recipient feels discomfort or any abnormalities, the recipient is moved to a first-aid room and medically checked by a medical doctor. After the observation period, the recipient can leave the observation room. Centre staff confirm the appropriate completion of the observation period.

**First-aid rooms in the centre**

The medical doctor medically checked each recipient, and the clinical findings were recorded. During May 24 to Sep 24, the average of recipients who visited first-aid rooms per day was 24 recipients. If needed, the recipients were treated by medicine and transported to neighbourhood hospitals. Information on outcome of transported recipients in neighbourhood hospitals are mailed to first-aid rooms and recorded.

### Supplementary method 2. The standard criteria to classify acute adverse events following immunisation

Acute adverse events following immunisation were classified according to the following standard criteria: (1) coincidental events, traumatic injuries, or symptoms/signs which have occurred before the recipient received the vaccine; (2) localised pain by vaccine product-related reactions, localised pain at the injection site without other symptoms/signs; (3) IHSR – one or more of following symptoms/signs were exhibited: urticarial or any type of rash, angioedema, local or generalised pruritus, wheezing, stridor, persistent cough, hoarseness, and anaphylaxis; (4) ISRR – symptoms/signs which were not accompanied by coincidental events, localised pain by vaccine product-related reactions or IHSR; discomfort in mouth and throat, palpitations, cold sweat, shortness of breath, chest pain, abdominal pain, nausea, vomiting, diarrhoea, vertigo, syncope, general weakness, numbness or loss of sensation, headache, malaise, hyperventilation/panic attack, photophobia, feeling of a hot flush, and vital signs abnormalities indicative of a vasovagal reflex (hypotension and/or bradycardia).

### Supplementary method 3. Sample size calculation

In Japanese population, the prevalence rate of variable of interests was estimated from 1% (malignancy) to 30·6% (hypertension) according to the Ministry of Health, Labour and Welfare, Japan (https://www.mhlw.go.jp/toukei_hakusho/toukei/). In this matched controls study, we determined the minimum sample size as 408 cases and 1,632 controls to detect minimum odds ratio of 3·0 for increased risk factors with immediate hypersensitivity reactions (IHSR) and immunisation stress-related response (ISRR) with 1% prevalence rate, 80% power and 0·05 two-sided type I error rate (α).

Before our study was started, Blumenthal and colleagues reported the incidence rate of IHSR to mRNA vaccine was 2·1% in vaccine recipients at first dose. For other drugs such as penicillin and contrast media, the incidence rate of IHSR was approximately 0·1%. The incidence rate of ISRR was considered as higher than IHSR. The target number of vaccination dose per day was set to 10,000 per day in the centre. Thus, we assumed that approximately 1,200-25,200 events of IHSR and more events of ISRR would be collected among 1,200,000 vaccine administration doses during the study period (May 24 to Sep 24).

However, the incidence rate of IHSR in this study was 0·03% and lower than a previous study and other drugs. Therefore, we collected only 318 events in 306 patients with IHSR in this study period. This sample size could detect minimum odds ratio of 5·0, 4·0, 3·0, and 2·0 for 0·5%, 1%, 2% and 5% prevalence rate of variable interests, respectively, with 80% power and 0·05 two-sided type I error rate (α).

| **Prevalence rate** | **Minimum odds ratio of detection** | **Power** | **Two-sided type I error rate (α)** | **Cases** | **Controls (1:4)** |
|---|---|---|---|---|---|
| 0·5% | 5 | 0·8 | 0·05 | 262 | 1048 |
| 0·5% | 4 | 0·8 | 0·05 | 413 | 1652 |
| 0·5% | 3 | 0·8 | 0·05 | 802 | 3208 |
| 0·5% | 2 | 0·8 | 0·05 | 2643 | 10572 |
| 1% | 4.0 | 0·8 | 0·05 | 211 | 844 |
| 1% | 3.0 | 0·8 | 0·05 | 408 | 1632 |
| 1% | 2.0 | 0·8 | 0·05 | 1136 | 5344 |
| 2% | 4.0 | 0·8 | 0·05 | 110 | 440 |
| 2% | 3.0 | 0·8 | 0·05 | 211 | 844 |
| 2% | 2.0 | 0·8 | 0·05 | 683 | 2732 |
| 5% | 4.0 | 0·8 | 0·05 | 50 | 200 |
| 5% | 3.0 | 0·8 | 0·05 | 93 | 372 |
| 5% | 2.0 | 0·8 | 0·05 | 292 | 1168 |

**Supplementary table 1. Case definition of immediate hypersensitivity reaction in sensitivity analyses**

| | Initial | IHSR-2 | IHSR-3 |
|---|---|---|---|
| **Symptoms/signs** | | | |
| **Any type of rash** | Included | Included | Included |
| **Angioedema** | Included | Included | Included |
| **Pruritus** | Included | Included | |
| **Wheezing** | Included | Included | Included |
| **Stridor** | Included | Included | Included |
| **Persistent cough** | Included | | |
| **Hoarseness** | Included | | |
| **Anaphylaxis** | Included | Included | Included |

IHSR: Immediate hypersensitivity reaction.

IHSR-2: Initial case definition without respiratory symptoms

IHSR-3: Initial case definition included only clinical signs

**Supplementary table 2. Case definition of immunisation stress-related response in sensitivity analyses**

| | Initial | IHSR-2 | IHSR-3 |
|---|---|---|---|
| **Symptoms/signs** | | | |
| **Mouth and throat discomfort** | Included | Included | |
| **Palpitations** | Included | Included | Included |
| **Cold sweat** | Included | Included | Included |
| **Shortness of breathing** | Included | Included | |
| **Chest pain** | Included | Included | |
| **Abdominal pain** | Included | | |
| **Nausea** | Included | | |
| **Vomiting** | Included | | |
| **Diarrhoea** | Included | | |
| **Vertigo** | Included | Included | Included |
| **Syncope** | Included | Included | Included |
| **General weakness** | Included | Included | Included |
| **Numbness or loss of sensation** | Included | Included | Included |
| **Headache** | Included | Included | Included |
| **Malaise** | Included | Included | Included |
| **Hyperventilation/panic attack** | Included | Included | Included |
| **Photophobia** | Included | Included | Included |
| **Feeling of hot flush** | Included | Included | Included |
| **Vasovagal reflex** | Included | Included | Included |

ISRR: Immunisation stress-related response

ISRR-2: Initial case definition of ISRR without gastrointestinal symptoms

ISRR-3: Initial case definition of ISRR without gastrointestinal and respiratory symptoms

Vasovagal reflex included hypotension and/or bradycardia

**Supplementary table 3. Participant of immediate hypersensitivity reactions groups in sensitivity analysis**

| | IHSR-2 | | | IHSR-3 | | |
|---|---|---|---|---|---|---|
| | Cases N=284 | Controls N=1,136 | *p* value | Cases N=188 | Controls N=752 | *p* value |
| **Demographic characteristic** | | | | | | |
| **Age, years** | | | | | | |
| **> 65 years** | 103 (36·3) | 441 (38·8) | 0·853 | 74 (39·4) | 324 (43·1) | 0·610 |
| **51-65 years** | 63 (22·2) | 237 (20·9) | | 42 (22·3) | 147 (19·5) | |
| **36-50 years** | 62 (21·8) | 249 (21·9) | | 36 (19·1) | 156 (20·7) | |
| **≤ 35 years** | 56 (19·7) | 209 (18·4) | | 36 (19·1) | 125 (16·6) | |
| **Sex** | | | | | | |
| **Male** | 61 (21·5) | 651 (57·3) | <0·001 | 44 (23·4) | 446 (59·3) | <0·001 |
| **Female** | 223 (78·5) | 485 (42·7) | | 144 (76·6) | 306 (40·7) | |
| **Comorbidities** | | | | | | |
| **Hypertension** | 40 (14·1) | 148 (13·0) | 0·626 | 33 (17·6) | 120 (16) | 0·582 |
| **Diabetes** | 12 (4·2) | 58 (5·1) | 0·646 | 8 (11·7) | 42 (5·9) | 0·587 |
| **Dyslipidaemia** | 16 (5·6) | 54 (4·8) | 0·541 | 15 (4·3) | 44 (5·6) | 0·312 |
| **Cardiovascular diseases** | 4 (1·4) | 39 (3·4) | 0·082 | 3 (1·6) | 30 (4·0) | 0·125 |
| **Asthma** | 20 (7·0) | 16 (1·4) | <0·001 | 14 (7·4) | 13 (1·7) | <0·001 |
| **Atopic dermatitis** | 4 (1·4) | 3 (0·3) | 0·033 | 4 (2·1) | 3 (0·4) | 0·033 |
| **Thyroid diseases** | 14 (4·9) | 11 (1·0) | <0·001 | 11 (5·9) | 9 (1·2) | <0·001 |
| **Malignancy** | 5 (1·8) | 19 (1·7) | 1·000 | 3 (1·6) | 16 (2·1) | 0·780 |
| **Mental disorders** | 6 (2·1) | 19 (1·7) | 0·615 | 4 (2·1) | 18 (2·4) | 1·000 |
| **History** | | | | | | |
| **Allergic episodes for drugs** | 81 (28·5) | 36 (3·2) | <0·001 | 54 (28·7) | 28 (3·7) | <0·001 |
| **Allergic episodes for foods** | 76 (26·8) | 29 (2·6) | <0·001 | 49 (26·1) | 20 (2·7) | <0·001 |
| **Vasovagal episode** | 8 (2·8) | 10 (0·9) | 0·016 | 3 (1·6) | 5 (0·7) | 0·202 |
| **Number of vaccine received** | | | | | | |
| **First** | 189 (66·5) | 756 (66·5) | 1·000 | 128 (68·1) | 512 (68·1) | 1·000 |
| **Second** | 95 (33·5) | 380 (33·5) | | 60 (31·9) | 240 (31·9) | |
| **Period** | | | | | | |
| **May 24-June 23** | 90 (31·7) | 360 (31·7) | 1·000 | 72 (38·3) | 288 (38·3) | 1·000 |
| **June 24-July 23** | 57 (20·1) | 228 (20·1) | | 30 (16·0) | 120 (16·0) | |
| **July 24-Aug 23** | 80 (28·2) | 320 (28·2) | | 50 (26·6) | 200 (26·6) | |
| **Aug 24-Sep 24** | 57 (20·1) | 228 (20·1) | | 36 (19·1) | 144 (19·1) | |

Data are n (%).

**Supplementary table 4. Participant of immunisation stress-related response groups in sensitivity analysis**

| | ISRR-2 Cases N=2,302 | ISRR-2 Controls N=9,208 | *p* value | ISRR-3 Cases N=2,129 | ISRR-3 Controls N=8,516 | *p* value |
|---|---|---|---|---|---|---|
| **Demographic characteristic** | | | | | | |
| **Age, years** | | | | | | |
| **> 65 years** | 400 (17·4) | 2200 (23·9) | <0·001 | 364 (17·1) | 1987 (23·3) | <0·001 |
| **51-65 years** | 342 (14·9) | 1784 (19·4) | | 312 (14·7) | 1657 (19·5) | |
| **36-50 years** | 576 (25·0) | 2596 (28·2) | | 516 (24·2) | 2424 (28·5) | |
| **≤ 35 years** | 984 (42·7) | 2628 (28·5) | | 937 (44·0) | 2448 (28·7) | |
| **Sex** | | | | | | |
| **Male** | 767 (33·3) | 5081 (55·2) | <0·001 | 727 (34·1) | 4699 (55·2) | <0·001 |
| **Female** | 1535 (66·7) | 4127 (44·8) | | 1402 (65·9) | 3817 (44·8) | |
| **Comorbidities** | | | | | | |
| **Hypertension** | 149 (6·5) | 850 (9·2) | <0·001 | 135 (6·3) | 753 (8·8) | <0·001 |
| **Diabetes** | 50 (2·2) | 326 (3·5) | 0·001 | 44 (2·1) | 294 (3·5) | 0·001 |
| **Dyslipidaemia** | 85 (3·7) | 383 (4·2) | 0·311 | 74 (3·5) | 348 (4·1) | 0·560 |
| **Cardiovascular diseases** | 51 (2·2) | 184 (2·0) | 0·510 | 45 (2·1) | 172 (2·0) | 0·784 |
| **Asthma** | 85 (3·7) | 112 (1·2) | <0·001 | 75 (3·5) | 106 (1·2) | <0·001 |
| **Atopic dermatitis** | 18 (0·8) | 33 (0·4) | 0·008 | 17 (0·8) | 30 (0·4) | 0·007 |
| **Thyroid diseases** | 49 (2·1) | 84 (0·9) | <0·001 | 43 (2·0) | 79 (0·9) | <0·001 |
| **Malignancy** | 25 (1·1) | 85 (0·9) | 0·473 | 21 (1·0) | 68 (0·8) | 0·395 |
| **Mental disorders** | 104 (4·5) | 119 (1·3) | <0·001 | 96 (4·5) | 107 (1·3) | <0·001 |
| **History** | | | | | | |
| **Allergic episodes for drugs** | 302 (13·1) | 285 (3·1) | <0·001 | 275 (12·9) | 260 (3·1) | <0·001 |
| **Allergic episodes for foods** | 286 (12·4) | 304 (3·3) | <0·001 | 254 (11·9) | 283 (3·3) | <0·001 |
| **Vasovagal episode** | 242 (10·5) | 129 (1·4) | <0·001 | 232 (10·9) | 121 (1·4) | <0·001 |
| **Number of vaccine received** | | | | | | |
| **First** | 1708 (74·2) | 6832 (74·2) | 1·000 | 1591 (74·7) | 6364 (74·7) | 1·000 |
| **Second** | 594 (25·8) | 2376 (25·8) | | 538 (25·3) | 2152 (25·3) | |
| **Period** | | | | | | |
| **May 24-June 23** | 449 (19·5) | 1796 (19·5) | 1·000 | 405 (19·0) | 1620 (19·0) | 1·000 |
| **June 24-July 23** | 305 (13·2) | 1220 (13·2) | | 275 (12·9) | 1100 (12·9) | |
| **July 24-Aug 23** | 998 (43·4) | 3992 (43·4) | | 941 (44·2) | 3764 (44·2) | |
| **Aug 24-Sep 24** | 550 (23·9) | 2200 (23·9) | | 508 (23·9) | 2032 (23·9) | |

Data are n (%).

**Supplementary table 5. Estimated incidence rates of adverse events following immunisation**

| | Total dose (N= 1,201,688) | | First doses (N= 611,779) | | Second doses (N= 589,909) | | |
|---|---|---|---|---|---|---|---|
| | **Total No. of events** | **Incidence rate (95% CI)** | **Total No. of events** | **Incidence rate (95% CI)** | **Total No. of events** | **Incidence rate (95% CI)** | ***p* value** |
| **IHSR** | 318 | 265·6 (236·3-295·3) | 213 | 348·2 (303·0-398·2) | 105 | 178·0 (145·6-215·5) | <0.001 |
| **ISRR** | 2558 | 2128·7 (2047·1-2212·7) | 1842 | 3010·9 (2875·1-3151·4) | 714 | 1210·4 (1123·2-1302·4) | <0.001 |
| **Anaphylaxis** | 2 | 1·7 (0·2-6·0) | 1 | 1·6 (0-9·1) | 1 | 1·7 (0-9·4) | 1.000 |
| **Vasovagal syncope** | 86 | 71·6 (57·2-88·4) | 76 | 123 (97·9-155·5) | 10 | 17.0 (8·1-31·1) | <0.001 |

Incidence rates of acute adverse events were calculated by using vaccine administration of doses in a centre as the denominator. Incidence rates were shown as per million doses. 95% CI; 95% confidence interval, IHSR; immediate hypersensitivity reactions, and ISRR; immunisation stress-related responses.

### Supplementary table 6. Number of events of clinical symptoms and signs

| | Total events | Immediate hypersensitivity reactions | Immunisation stress-related responses |
|---|---|---|---|
| **Skin, facial and oral symptoms/signs** | | | |
| **Any type of rash** | 193 | 193 | 0 |
| **Angioedema** | 7 | 7 | 0 |
| **Pruritus** | 147 | 147 | 0 |
| **Mouth and throat discomfort** | 179 | 33 | 146 |
| **Cardiovascular symptoms/signs** | | | |
| **Palpitations** | 298 | 8 | 290 |
| **Cold sweat** | 125 | 1 | 124 |
| **Respiratory symptoms/signs** | | | |
| **Wheezes or stridor** | 4 | 4 | 0 |
| **Hoarseness** | 1 | 1 | 0 |
| **Persistent cough** | 25 | 25 | 0 |
| **Shortness of breathing** | 138 | 13 | 125 |
| **Chest pain** | 37 | 0 | 37 |
| **Gastrointestinal symptoms/signs** | | | |
| **Abdominal pain** | 21 | 3 | 18 |
| **Nausea** | 281 | 12 | 269 |
| **Vomiting** | 14 | 0 | 14 |
| **Diarrhoea** | 2 | 0 | 2 |
| **Neurological symptoms/signs** | | | |
| **Vertigo** | 891 | 8 | 883 |
| **Syncope** | 86 | 0 | 86 |
| **General weakness** | 47 | 2 | 45 |
| **Numbness or loss of sensation** | 411 | 8 | 403 |
| **Other symptoms/signs** | | | |
| **Headache** | 112 | 3 | 109 |
| **Malaise** | 497 | 11 | 486 |
| **Hyperventilation/Panic attack** | 30 | 1 | 29 |
| **Photophobia** | 28 | 3 | 25 |
| **Feeling of hot flush** | 22 | 0 | 22 |
| **Abnormalities of vital signs** | | | |
| **Hypotension** | 458 | 6 | 452 |

| | | | |
|---|---|---|---|
| **Hypertension** | 171 | 32 | 139 |
| **Bradycardia** | 385 | 10 | 375 |
| **Tachycardia** | 33 | 6 | 27 |
| **Tachypnoea** | 40 | 3 | 37 |

Data are n. Hypotension; systolic blood pressure < 90 mmHg and/or diastolic blood pressure < 60 mmHg, hypertension; systolic blood pressure > 180 mmHg and/or diastolic blood pressure > 110 mmHg, bradycardia; heart rate < 60 beat per minutes, tachycardia; heart rate > 120 beat per minutes and tachypnoea; respiratory rate >24 per min.

**Supplementary table 7. Incidence rate of clinical symptoms and signs**

| | Total events | Immediate hypersensitivity reactions | Immunisation stress-related responses |
|---|---|---|---|
| **Skin, facial and oral symptoms/signs** | | | |
| **Any type of rash** | 160·6 (138·8-184·9) | 160·6 (138·8-184·9) | 0 |
| **Angioedema** | 5·8 (2·3-12·0) | 5·8 (2·3-12·0) | 0 |
| **Pruritus** | 122·3 (103·4-143·8) | 122·3 (103·4-143·8) | 0 |
| **Mouth and throat discomfort** | 149·0 (127·9-172·4) | 27·5 (18·9-38·6) | 121·5 (102·6-142·9) |
| **Cardiovascular symptoms/signs** | | | |
| **Palpitations** | 248·0 (220·6-277·8) | 6·7 (2·9-13·1) | 241·3 (214·4-270·8) |
| **Cold sweat** | 104·0 (86·6-123·9) | 0·8 (0-4·6) | 103·2 (85·8-123·0) |
| **Respiratory symptoms/signs** | | | |
| **Wheezes or stridor** | 3·3 (0·9-8·5) | 3·3 (0·9-8·5) | 0 |
| **Hoarseness** | 0·8 (0-4·6) | 0·8 (0-4·6) | 0 |
| **Persistent cough** | 20·8 (13·5-30·7) | 20·8 (13·5-30·7) | 0 |
| **Shortness of breathing** | 114·8 (96·5-135·7) | 10·8 (5·8-18·5) | 104·0 (86·6-123·9) |
| **Chest pain** | 30·8 (21·7-42·4) | 0 | 30·8 (21·7-42·4) |
| **Gastrointestinal symptoms/signs** | | | |
| **Abdominal pain** | 17·5 (10·8-26·7) | 2·5 (0·5-7·3) | 15·0 (8·9-23·7) |
| **Nausea** | 233·8 (207·3-262·8) | 10·0 (5·2-17·4) | 223·9 (197·9-252·3) |
| **Vomiting** | 11·7 (6·4-19·5) | 0 | 11·7 (6·4-19·5) |
| **Diarrhoea** | 1·7 (0·2-6·0) | 0 | 1·7 (0·2-6) |
| **Neurological symptoms/signs** | | | |
| **Vertigo** | 741·5 (693·6-791·8) | 6·7 (2·9-13·1) | 734·8 (687·2-784·9) |
| **Syncope** | 71·6 (57·2-88·4) | 0 | 71·6 (57·2-88·4) |
| **General weakness** | 39·1 (28·7-52·0) | 1·7 (0·2-6) | 37·4 (27·3-50·1) |
| **Numbness or loss of sensation** | 342·0 (309·8-376·7) | 6·7 (2·9-13·1) | 335·4 (303·4-369·8) |
| **Other symptoms/signs** | | | |
| **Headache** | 93·2 (76·7-112·1) | 2·5 (0·5-7·3) | 90·7 (74·5-109·4) |
| **Malaise** | 413·6 (378-451·6) | 9·2 (4·6-16·4) | 404·4 (369·3-442) |
| **Hyperventilation/Panic attack** | 25·0 (16·8-35·6) | 0·8 (0-4·6) | 24·1 (16·2-34·7) |
| **Photophobia** | 23·3 (15·5-33·7) | 2·5 (0·5-7·3) | 20·8 (13·5-30·7) |
| **Feeling of hot flush** | 18·3 (11·5-27·7) | 0 | 18·3 (11·5-27·7) |
| **Abnormalities of vital signs** | | | |
| **Hypotension** | 381·1 (347-417·7) | 5·0 (1·8-10·9) | 376·1 (342·3-412·5) |

| | | | |
|---|---|---|---|
| **Hypertension** | 142·3 (121·8-165·3) | 26·6 (18·2-37·6) | 115·7 (97·2-136·6) |
| **Bradycardia (< 60 bpm)** | 320·4 (289·2-354·0) | 8·3 (4·0-15·3) | 312·1 (281·3-345·3) |
| **Tachycardia (>120 bpm)** | 27·5 (18·9-38·6) | 5·0 (1·8-10·9) | 22·5 (14·8-32·7) |
| **Tachypnoea (>24/min)** | 33·3 (23·8-45·3) | 2·5 (0·5-7·3) | 30·8 (21·7-42·4) |

Data are incidence rates per million doses (95% confidence interval). The incidence rates were estimated using vaccine doses administrated as the denominator. Hypotension; systolic blood pressure < 90 mmHg and/or diastolic blood pressure < 60 mmHg, hypertension; systolic blood pressure > 180 mmHg and/or diastolic blood pressure > 110 mmHg, bradycardia; heart rate < 60 beat per minutes, tachycardia; heart rate > 120 beat per minutes and tachypnoea; respiratory rate >24 per min.

**Supplementary figure 1. Flow diagram of immediate hypersensitivity reactions groups in sensitivity analysis**

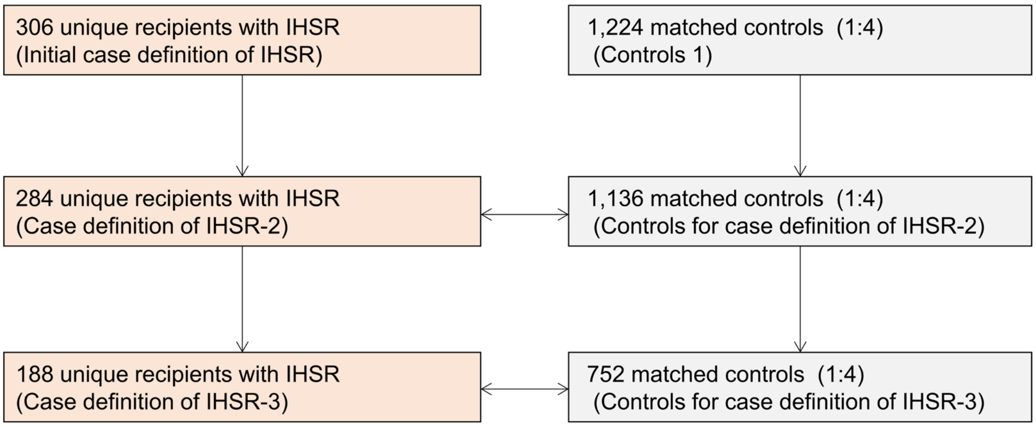


We excluded recipients with missing data in sensitivity analyses (complete case analysis).

**Supplementary figure 2. Flow diagram of immunisation stress-related response groups in sensitivity analysis**

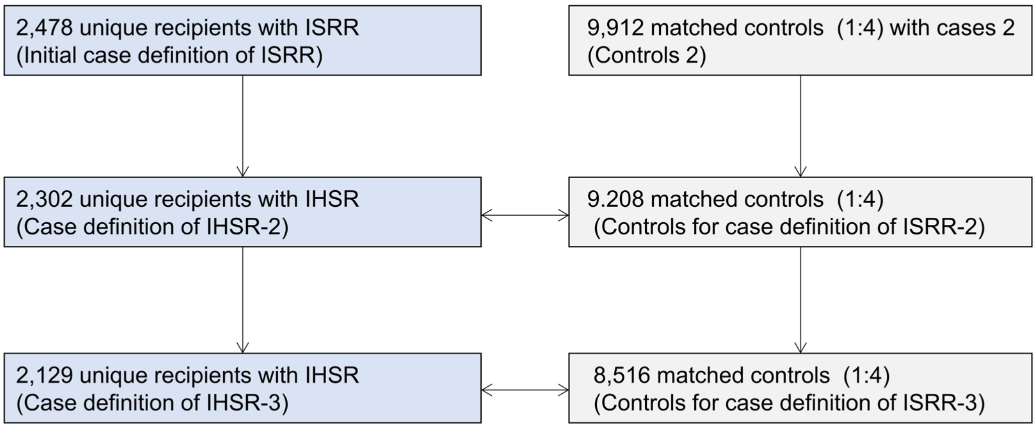


We excluded recipients with missing data in sensitivity analyses (complete case analysis).

**Supplementary figure 3. Multivariable conditional logistic regression for immediate hypersensitivity reactions group in sensitivity analysis**

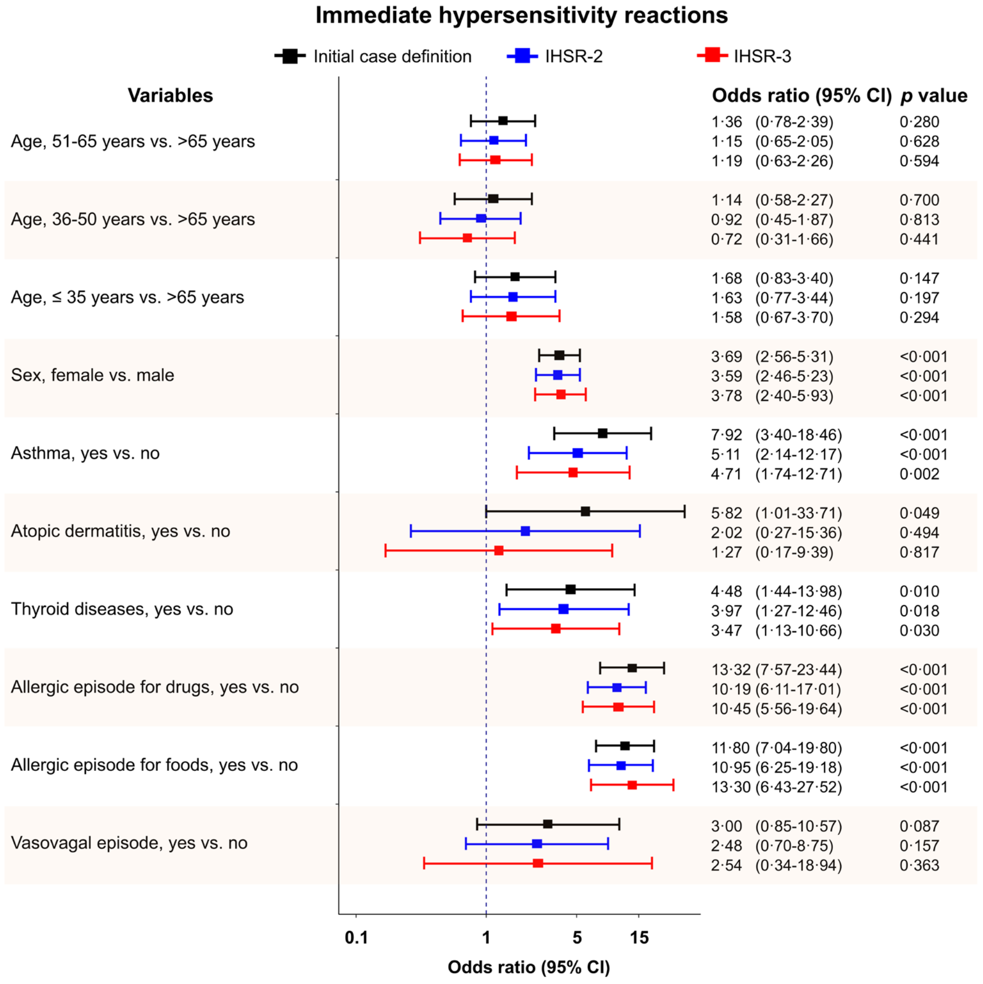


Forest plot showing the odds ratio for increased risk of immediate hypersensitivity reactions (IHSR) using multivariable analysis of conditional logistic regression analysis. Black, blue and red plots indicate estimated odds ratio by initial case-definition, case-definition of IHSR-2, and IHSR-3, respectively.

Black, blue and red horizontal lines indicate estimated odds ratio and 95% confidence intervals by initial case-definition, case-definition of IHSR-2, and IHSR-3, respectively.

**Supplementary figure 4. Multivariable conditional logistic regression for immunisation stress-related response group in sensitivity analysis**

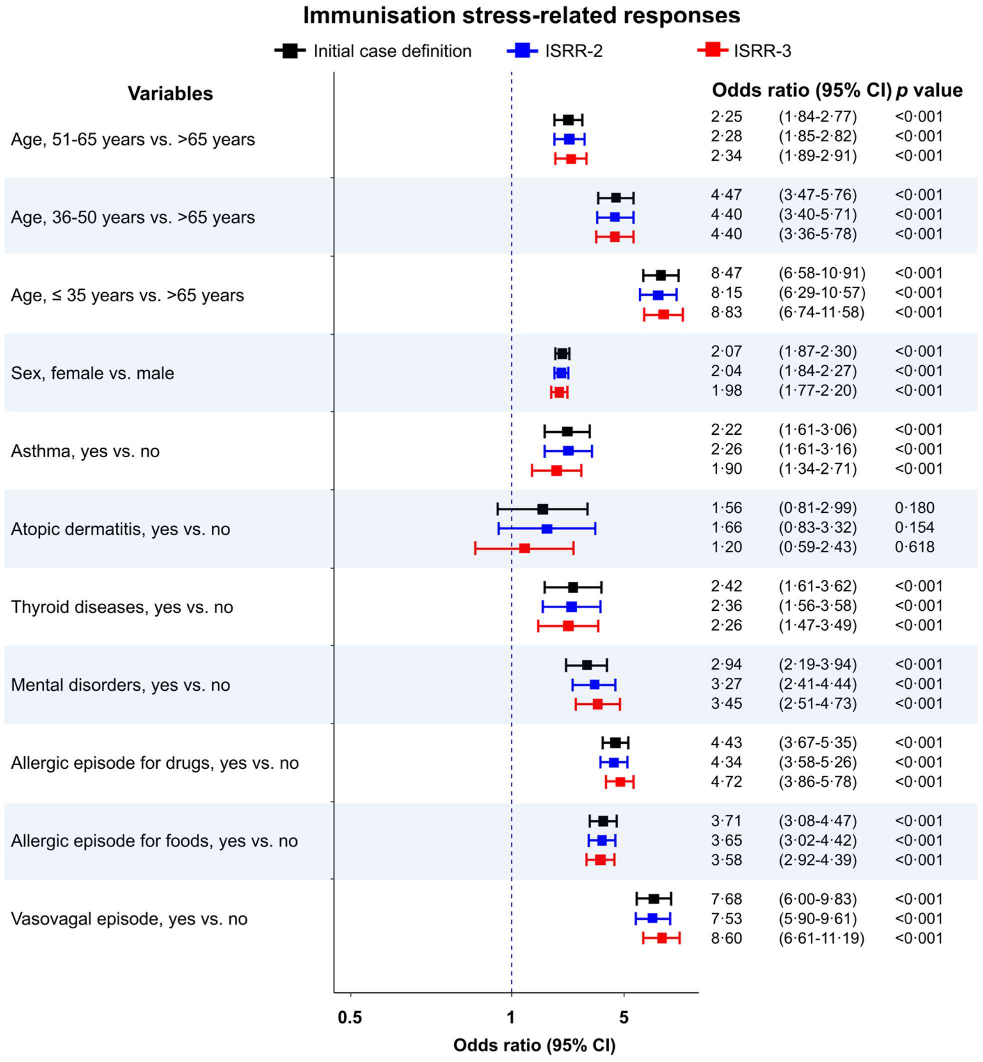


Forest plot showing the odds ratio for increased risk of immunisation stress-related responses (ISRR) using multivariable analysis of conditional logistic regression analysis. Black, blue and red plots indicate estimated odds ratio by initial case-definition, case-definition of ISRR-2, and ISRR-3, respectively.

Black, blue and red horizontal lines indicate estimated odds ratio and 95% confidence intervals by initial case-definition, case-definition of ISRR-2, and ISRR-3, respectively.
